# Investigating misclassification of type 1 diabetes in a population-based cohort of British Pakistanis and Bangladeshis using polygenic risk scores

**DOI:** 10.1101/2023.08.23.23294497

**Authors:** Timing Liu, Alagu Sankareswaran, Gordon Paterson, Genes & Health Research Team, Diane P Fraser, Sam Hodgson, Qin Qin Huang, Teng Hiang Heng, Meera Ladwa, Nick Thomas, David A. van Heel, Michael N. Weedon, Chittaranjan S Yajnik, Richard A Oram, Giriraj R Chandak, Hilary C Martin, Sarah Finer

## Abstract

**Aims:** Correct classification of type 1 (T1D) and type 2 diabetes (T2D) is challenging due to overlapping clinical features and the increasingly early onset of T2D, particularly in South Asians. We used polygenic risk scores (PRSs) in a British Bangladeshi and Pakistani population with diabetes to estimate the proportion and misclassification rate of T1D in insulin-treated individuals with ambiguous features.

**Methods:** Using linked health records from the Genes & Health cohort (n=38,344) we defined four groups: 31 T1D cases, 1,842 T2D cases, and after excluding these, 839 insulin-treated individuals with ambiguous features and 5,174 controls. Combining these with 307 confirmed T1D cases and 307 controls from India, we calculated ancestry-corrected PRSs for T1D and T2D, with which we estimated the proportion of T1D cases within the ambiguous group and evaluated misclassification.

**Results:** We estimated that the prevalence of T1D was ∼6% within the ambiguous group, or ∼4.5% within the subset who had T2D codes in their health records. We saw no significant association between the T1D or T2D PRS and BMI at diagnosis, time to insulin, or the presence of T1D or T2D diagnostic codes amongst the T2D or ambiguous cases, suggesting that these clinical features are not particularly helpful at aiding diagnosis in ambiguous cases.

**Conclusions:** We estimate that about one in twenty of British Pakistanis and Bangladeshis with diabetes who are treated with insulin and have ambiguous clinical features have been classified incorrectly in their health records, and in fact have T1D. This emphasises that robust identification of T1D cases and appropriate clinical care may require routine measurement of diabetes autoantibodies and C-peptide.

**Research in context:** *What is already known about this subject?:*

- Correct classification of type 1 (T1D) and type 2 diabetes (T2D) is challenging due to overlapping clinical features and the increasingly early onset of T2D, particularly in people of South Asian descent.
- Polygenic risk scores (PRSs) are useful tools to aid the classification of people with diabetes.

*What is the key question?:*

- What proportion of insulin-treated diabetic individuals with ambiguous clinical features have been clinically misclassified and in fact have T1D, amongst a cohort of British Pakistani and Bangladeshi adults?

*What are the new findings?:*

- Based on analyses of polygenic risk scores, the prevalence of T1D was found to be ∼6% within patients who were insulin-treated but with ambiguous features, and ∼4.5% were estimated to have been misclassified.
- Clinical features such as BMI at diagnosis, time to insulin, or presence of T1D/T2D codes were not significantly associated with T1D or T2D PRS.

*How might this impact on clinical practice in the foreseeable future?:*

- These findings emphasise the importance of routine collection of diabetes autoantibodies and C-peptide measurements to identify T1D cases robustly, especially in countries where diabetes cases are diagnosed in primary care without input from diabetologists.

---

Type 1 and type 2 diabetes (T1D and T2D respectively) are classified aetiologically, and the two conditions display a different disease course, treatment requirement and lived experience (1,2). For clinicians, correctly classifying T1D and T2D may be challenging due to overlapping clinical features, and the increasingly early onset of T2D. Rising obesity levels in children and young people may reduce the utility of adiposity measures (e.g. body mass index) in distinguishing between T1D and T2D cases. Additionally, a lack of recognition of late-onset T1D cases may lead to their incorrect classification as T2D (3).

Diabetes misclassification occurs when a clinician-recorded diagnosis of T1D or T2D does not match the true (aetiological) category to which the subject belongs (4), but it can be difficult to distinguish from erroneous miscoding in health records. Previous estimates of miscoding and misclassification suggest that up to 13% of diabetes cases recorded in British general practices are affected, but these studies do not establish a ground truth and rely on a complex algorithmic definition of type 1 and type 2 diabetes using clinical features and the use of oral versus insulin treatment (4–7). The correct classification of type 1 and type 2 diabetes is suggested by rapid requirement to insulin (within 3 years of diagnosis) (8). However, without specialised tests to estimate endogenous insulin secretion (serum C-peptide measurement) and/or to determine if diabetes-specific autoimmunity is present (measurements of diabetes autoantibodies), diabetes type cannot be conclusively determined. Additionally, even if diabetes autoantibodies are measured, the sensitivity and specificity of the measurement vary by age. Misclassification of diabetes may be represented by the presence of incorrect or multiple different diagnostic codes relating to diabetes in an individual’s electronic health record. However, the presence of multiple different diagnostic codes may simply reflect their erroneous use rather than necessarily revisions to diagnostic classification, making it difficult to correctly classify diabetes in real-world electronic health record datasets (9).

Misclassification rates of diabetes in people of South Asian descent are not known (10,11) but are expected to be greater than those in White Europeans, due to high prevalence of T2D and its greater tendency to present in people who are slim, young and in some cases have features of insulin deficiency (12,13). The high prevalence and early onset of T2D in South Asians may bias clinicians towards making diagnosis of T2D over T1D even when clinical features commonly considered supportive of a T1D diagnosis (e.g. requiring insulin soon after diagnosis) are present (8,14). Inaccurate classification of a true T1D case as T2D could result in severe acute harms including diabetic ketoacidosis (15,16) but could also lead to increased risk of long-term diabetic complications (15). Misclassification of a true T2D case and subsequent prescription of insulin may lead to harms including increased risk of hypoglycaemic events (17). Furthermore, patients who are misclassified as T1D may miss out on oral therapies that are highly effective at reducing the risk of T2D complications.

Large-scale genome-wide association studies (GWAS) have derived major insights into the genetic aetiology of complex diseases such as T1D and T2D (18–20) and have led to the construction of polygenic risk scores (PRS) that could have clinical benefit through use in risk prediction and characterisation of disease heterogeneity (3,21–24). Recent work has shown that PRS for T1D and T2D are useful tools to aid the classification of people with diabetes (13,22,25). Historically, most genomic studies are focused on individuals with European ancestry (26) and the extent to which their findings are transferable into diverse ancestry groups differs between target groups (27) and traits (28). Large multi-ancestry genome-wide association studies for T2D have recently been performed (18,29). To determine the potential of diabetes PRSs to aid disease classification and precision medicine, it is imperative that they are evaluated at scale and in understudied ancestry groups, particularly those in which misclassification is suspected.

In this study we therefore aim to apply genetic ancestry-optimised T1D and T2D PRSs (30) to a population of British South Asians with diabetes to estimate the proportion and misclassification of T1D in those with ambiguous features who are aged under 60 years and treated with insulin. Secondly, we aim to test whether T1D or T2D PRSs were associated with the clinical characteristics commonly used to determine diabetes type clinically. We used the Genes and Health (G&H) cohort based in East London, UK, which combines genomic and detailed electronic health record data for over 44,000 people of British Bangladeshi and Pakistani descent (31). The population from which the G&H subjects are recruited has a high risk of T2D, approximately twice that of the local White-European population (32). The combination of high-quality phenotypic and genetic data gives us a unique opportunity to study misclassification of diabetes in this understudied South Asian-ancestry population.

## Research Design and Methods

### G&H study population and clinical codes

The Genes & Health cohort has been described previously (21,31,33). Briefly, G&H is a community-based study which recruits British Pakistani and Bangladeshi individuals aged 16 years and older from National Health Service (NHS) and community settings in the UK. All volunteers consent to lifelong electronic health record (EHR) access and donate a saliva sample for genetic studies. G&H was approved by the London South East NRES Committee of the Health Research Authority (14/LO/1240).

For this study, we used the June 2021 data release, selecting 38,344 of the 46,132 volunteers recruited in east London (median age 43 yrs; 54.4% female; 45.6% male) who had primary health care record data available. Additional secondary care data was available for 22,713 of the 38,244 individuals who had any interactions with Barts Health NHS Trust, the largest secondary care trust in the area providing inpatient and outpatient services, including specialist diabetes care. Individuals were excluded (N=94) if they had registered at different GP practices with incongruent demographic data (specifically year of birth). Definitions of clinical diagnosis can be found in ESM Methods, with key codes given in ESM Tables 1 and 2. Quantitative clinical measures were cleaned as described in ESM Methods.

### Defining ambiguous cases, reference cases and controls in G&H

We filtered the available data to define four groups of mutually exclusive individuals, as follows. If electronic health records did not contain sufficient historic information to inform the T1D reference case selection (e.g. unavailability of historic prescribing information), additional case note review was undertaken by two experienced diabetologists (SF, ML).

1) T1D reference cases:

- Individuals with a clinical code for T1D or T2D. (In practice, as Table 1 shows, most of the thirty-one individuals ultimately included in this group did have a T1D code, but we also included seven who had only a T2D code because, based on review from two senior diabetologists, we believe the following criteria makes it highly likely they are truly T1D and this is a miscoding error.)
- AND had a time to insulin from diagnosis between 0.5 - 1 years
- AND (a) for those aged under 30 at diagnosis either insulin deficiency OR positive diabetes autoantibodies were required; (b) for those aged 30 to 60 years at diagnosis ONLY evidence of insulin deficiency was used. No individuals who were aged over 60 at diagnosis passed the above criteria.
2) T2D reference cases:

- Individuals with a clinical code for T2D
- AND duration of diabetes > 3 years
- AND had received oral anti-hyperglycaemic treatment but never insulin
- AND no confirmed insulin deficiency or autoantibody positivity
3) Ambiguous group

- Individuals with a clinical code for T1D or T2D
- Age at diagnosis 60 years old or younger
- AND had received an insulin prescription within the most recent year of data linkage
- AND did not meet criteria for T1D or T2D reference cases
4) Non-diabetic controls:

- Individuals with no diagnostic code ever recorded for: T1D, T2D, Secondary/rare diabetes and pancreatic disease/surgery, diabetes risk states (defined in ESM Table 3)
- AND has no confirmed insulin deficiency or autoantibody positivity
- AND has never been prescribed diabetes medications (any)

The number of individuals remaining after these filtering steps are shown in ESM Table 4.

Our stringent criteria to define confirmed T1D resulted in a group of only 31 individuals (Table 1), and therefore a separate T1D reference population was used from an Indian cohort, the details of which are presented below.

**Table 1:** Clinical characteristics recorded within one year of diagnosis of individuals in the ambiguous, T1D and T2D groups in G&H, and in the G&H non-diabetic controls. In the top part of the table, quantitative traits are presented as the median (quartiles), and in the bottom part, count variables are presented as sample size (percentage). Significance was assessed using Kruskal-Wallis tests (non-parametric one-way ANOVA). Dunn’s and Chi-squared post-hoc tests were performed where appropriate (ESM Table 5). * significant (p<0.05) vs T1D group; ^#^ significant (p<0.05) vs T2D group; ^$^ significant (p<0.05) vs ambiguous group. ^@^ Eighteen controls for whom both male and female genders were recorded in the health records were excluded from clinical comparisons.

| Characteristic | N. with data | Median value (quartiles) |  |  |  | p-value (ANOVA) |
| --- | --- | --- | --- | --- | --- | --- |
|  |  | Ambiguous<br>N = 839 | T1D<br>N = 31 | T2D<br>N = 1,842 | Controls<br>N = 5,174 |  |
| Age at diagnosis (years) | 2,712 | 40 (33, 46)<sup>*#</sup> | 23 (13, 34)<sup>\$#</sup> | 45 (39, 53)<sup>\$*</sup> | NA | <0.001 |
| BMI (kg/m<sup>2</sup>) | 5,928 | 27.8 (25.1, 31.8) | 26.4 (20.4, 31.8) | 28.4 (25.6, 32.1) | 26.3 (22.8, 29.0) | 0.08 |
| HDL (mmol/l) | 4,324 | 1.09 (0.90, 1.23) | 1.20 (1.10, 1.43) | 1.04 (0.90, 1.21) | 1.27 (1.05, 1.44) | 0.19 |
| Triglycerides (mmol/l) | 4,134 | 2.00 (1.37, 2.81) | 1.65 (1.05, 2.54) | 2.00 (1.40, 2.98) | 1.46 (0.85, 1.75) | 0.39 |
| HbA1c (mmol/mol) | 4,814 | 67 (55, 87)<sup>#</sup> | 78 (57, 111)<sup>#</sup> | 59 (51, 76)<sup>\$*</sup> | 36 (33, 38) | <0.001 |
| C-peptide (pmol/L) | 57 | 905 (564, 1228)<sup>*</sup> | 162 (21, 168)<sup>\$#</sup> | 1056 (880, 1179)<sup>*</sup> | NA | <0.001 |
| Time to insulin (years) | 864 | 10 (5, 15) | 5 (1, 12) | NA | NA | 0.025 |
|  |  | Number of individuals (%) |  |  |  |  |
| Gender | 7,868 |  |  |  |  | <0.001 |
| Female | | 450 (54%)<sup>#</sup> | 12 (39%) | 749 (41%)<sup>\$</sup> | 2,962 (57%)<sup>@</sup> | |
| Male | | 389 (46%)<sup>#</sup> | 19 (61%) | 1,093 (59%)<sup>\$</sup> | 2,194 (42%)<sup>@</sup> | |
| Autoimmune Condition Present (n, %) | 7,886 | 43 (5.1%)<sup>#</sup> | 3 (9.7%) | 53 (2.9%)<sup>\$</sup> | 117 (2.2%) | 0.003 |
| Only T1D Code (n, %) | 7,886 | 18 (2%)<sup>*</sup> | 15 (48%)<sup>\$</sup> | 0 (0%) | 0 (0%) | <0.001 |
| Only T2D Code (n, %) | 7,886 | 18 (2%)<sup>*</sup> | 7 (23%)<sup>\$#</sup> | 1,822 (99%)<sup>\$*</sup> | 0 (0%) | <0.001 |
| T1D & T2D Code (n, %) | 7,886 | 105 (13%)<sup>*#</sup> | 9 (29%)<sup>\$#</sup> | 20 (1%)<sup>\$*</sup> | 0 (0%) | <0.001 |
| Diabetes Auto-antibodies tested (n) | 69 | 34 (4%) | 15 (48%) | 20 (1%) | 0 (0%) | <0.001 |
| Positive (n, %) | | 9 (26%)<sup>*#</sup> | 11 (73%)<sup>\$#</sup> | 0 (0%)<sup>\$*</sup> | 0 (0%) | |
| Negative (n, %) |  | 25 (74%) | 4 (27%) | 20 (100%) | 0 (0%) |  |

### Statistical analysis of clinical data

For comparisons between baseline groups, Kruskall-Wallis tests were performed with Dunn’s post-hoc where appropriate. For comparisons of two groups, student’s t-tests were used to compare continuous measures and chi-squared tests were used to compare binary end-points.

### Preparation of G&H genotype data

We used the June 2021 data freeze which included 46,132 individuals genotyped on the Illumina Infinium Global Screening Array v3 chip (GRCh38). Quality control, inference of genetic ancestry and inference of relatedness are described in the ESM Methods. After quality control, restriction to individuals fulfilling the clinical criteria described above, and removal of relatives, we retained 7,886 unrelated individuals with genetically-inferred Pakistani or Bangladeshi ancestry. These comprised 31 T1D cases, 1,842 T2D cases, 839 ambiguous cases and 5,174 non-diabetic controls. We combined genetic data from these individuals with the Indian cohort described below.

### The Indian cohort of T1D cases and controls

The Indian cohort consisted of 332 T1D cases and 317 non-diabetic controls who were recruited in Pune, India via detailed phenotypic characterisation and robust diagnostic classification, using the same methods as those described in Harrison et al. 2020 (30). The Institutional Ethics Committee of the KEM Hospital Research Centre, Pune, India (KEMHRC ID No1737 & KEMHRC ID No PhD19) approved the study, and all methods were performed in accordance with the relevant guidelines and regulations. Individuals were genotyped on the Illumina GSA-24v3 chip. Quality control of the genotype data is described in the ESM Methods. After removing principal component outliers and third-degree relatives or closer using PropIBD metric from KING, 307 cases and 307 controls remained in the dataset.

### Principal component analysis and imputation of G&H and Indian samples

We combined the genetic data from the 7,886 unrelated G&H individuals and the 614 unrelated Indian individuals to perform a principal component analysis (PCA) which we subsequently used to correct the T1D and T2D polygenic risk scores (PRSs) for genetic ancestry differences (described below). The preparation of this combined dataset, the PCA, and imputation to TOPMED r2 are described in ESM Methods.

### PRS calculation and PC correction

For type 1 diabetes, we used a previously-published PRS which included ten SNPs including two that, in combination, tag HLA-DR3/DR4-DQ8 haplotype. This PRS had been shown to be reasonably transferable to the Indian population (30,34). Oram et al. showed that this 10-SNP score performed almost as well as the larger 30-SNP score (34).

For type 2 diabetes, the PRS was calculated using PRSice2.2 using SNPs that had *P*<1×10^-4^ in the largest recent trans-ancestry meta-analysis (36), as we found that the area under the receiver operating curve was the highest at this *P* value cut-off. Clumping R^2^ was set to 0.1, and European samples from 1000 Genomes Project were used as the LD reference.

We observed significant differences in the PRS distributions of non-diabetic controls between the genetically-inferred ancestry groups (ESM Figure 1), which are well known to exist due to differences in demographic history (35). We thus decided to correct these by regressing out the principal components (PCs). A scree plot (ESM Figure 2) suggested that five PCs were sufficient to explain most of the variation (ESM Figure 3). Since ancestry was correlated with case status within our cohort (i.e. the majority of T1D cases were Indian and the T2D cases were Pakistani/Bangladeshi), we regressed the PRSs on these five PCs in the controls only for both the T1D and T2D PRSs:

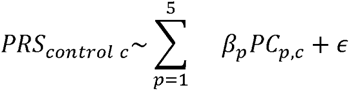

We then used the estimated *β*s for each PC to calculate the expected PRS for each case and control individual *i* as follows:

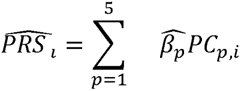

We then calculated the residuals which we used in subsequent analyses:

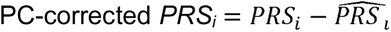

### Statistical analysis of PRSs

We employed the approach from Evans et al. (22) to estimate the prevalence of T1D in the set of ambiguous cases from G&H (*f_T1D_*). We did this using three approaches. In all cases we used the PC-corrected PRSs constructed as described above. In the first approach, we used the small sample of G&H T1D cases as “true cases” and the G&H T2D cases as “non-cases” to estimate *f_T1D_* using the PC-corrected T1D PRS. Since we had only a small sample of clear T1D cases in G&H, in the second approach we used the larger sample of Indian T1D cases as our “true cases” and the G&H T2D cases as “non-cases”, and used the PC-corrected T1D PRSs. In the third approach, we used the T2D cases and non-diabetic controls from G&H to estimate the fraction of the ambiguous cases that had T2D (*f_T2D_*) using the PC-corrected T2D PRS, and then calculated *f_T1D_* as 1-*f_T2D_.* With each approach, we used the three statistical methods from (22): the means method, the Earth Mover’s Distance (EMD), and the Kernel Density Estimation (KDE) method.

### Association between PRS and clinical parameters

We selected three clinical parameters routinely used by clinicians and researchers to classify diabetes diagnosis: age at diagnosis, BMI at diagnosis and time to insulin start (3,36). We used multiple linear regression to assess the association between the T1D or T2D PRS and these clinical parameters within either the ambiguous cases or T2D cases (Figure 3). Analysis was performed using R 4.0.1 and Python 3.8.

## Results

### Definition and clinical characteristics of T1D, T2D and ambiguous cases in G&H

Clinical data were available for a total of 38,344 individuals. ESM Table 4 indicates the results of our filtering process to define T1D, T2D, ambiguous cases and nondiabetic controls, who were subsequently filtered to remove related individuals. Using this process, we were able to identify 31 T1D reference cases, 1,842 T2D reference cases, 839 further diabetes cases treated with insulin and deemed ambiguous by the criteria outlined above, and 5,174 nondiabetic controls. The majority of the individuals we removed were excluded due to having diabetes risk states (e.g. ‘at risk of diabetes mellitus’ or ‘family history of diabetes mellitus’) in their health records, or due to being putative T2D cases not on oral antihyperglycemic agents. Using ANOVA, we compared clinical features between the groups (Table 1) and found no significant difference in BMI at diagnosis. There were significant differences across groups in age and HbA1c at diagnosis, C-peptide (ANOVA *P*<0.001) and diabetes autoantibody positivity (ANOVA *P*<0.01), consistent with clinical expectation. However, C-peptide and diabetes autoantibodies were rarely measured (0.7% and 2.5% of diabetes cases, respectively). The ambiguous group displayed intermediate clinical features, with post-hoc pairwise comparisons (ESM Table 5) demonstrating differences with both T1D and T2D reference groups in age at diagnosis (40 years for the ambiguous group versus 23 yrs for T1D cases and 45 yrs for T2D cases), C-peptide significantly higher than the T1D but not T2D reference cases (905 for the ambiguous vs 162 for T1D vs 1056 pmol/l for T2D groups, respectively). In contrast, HbA1c at diagnosis was significantly higher in the ambiguous group (67 mmol/mol) than T2D (59 mmol/mol) but not T1D (78 mmol/mol) cases. Importantly, clinical codes did not differentiate these groups reliably, with 39% of T1D cases having a T2D code present in their electronic health records, and only 77% actually having a T1D code. The ambiguous group was predominantly coded as having T2D: 93% had only a T2D code, but 7% had a T1D code present either with or without a T2D code.

### Estimating the fraction of T1D cases using PRSs

Evans *et al.* recently proposed a mathematical framework for estimating the prevalence of a given disease within a cohort using PRSs (22). This relies on estimating the proportion of true cases and non-cases from the PRS distribution of a sample of individuals that contains a mixture of these. We wanted to use this framework to estimate the proportion of T1D within the set of 839 ambiguous cases. However, since we only had 31 definite T1D cases from G&H, we anticipated this would be insufficient to produce an accurate estimate of the prevalence of T1D within the ambiguous group. We thus combined our data with a larger set of 307 cases and 307 controls from Pune, India, and regressed out genetic principal components from the T1D and T2D PRSs to correct for ancestry (ESM Figure 3). PC-corrected PRSs did not show significant differences between the different ancestry groups amongst the controls (ESM Figure 1b and 4b). We then applied three approaches to estimate the prevalence of T1D within the ambiguous group using the PC-corrected PRSs, and for each, used three different statistical methods to estimate the mixture proportion (see Methods).

We found that the three approaches produced very similar estimates of the fraction of T1D cases in our ambiguous group, with the point estimates ranging from 3.6% to 10.2% (median 5.9%) and confidence intervals from 0% to 15.2% (Figure 1a). Estimates were also very similar between three different statistical approaches used to estimate the mixture proportion, with confidence intervals all overlapping. We then performed the same analysis removing individuals (n = 18) from the ambiguous group who only had a T1D code present in their clinical records, considering that they were more likely to be true T1D diagnoses. This therefore gave an estimate of the proportion of individuals who are likely to be misclassified. The point estimates of prevalence (and presumed misclassification estimates) ranged from 1.9% to 7.8%, with a median of 4.5%, and confidence intervals from 0% to 12.6% (Figure 1b).

**Figure 1.**
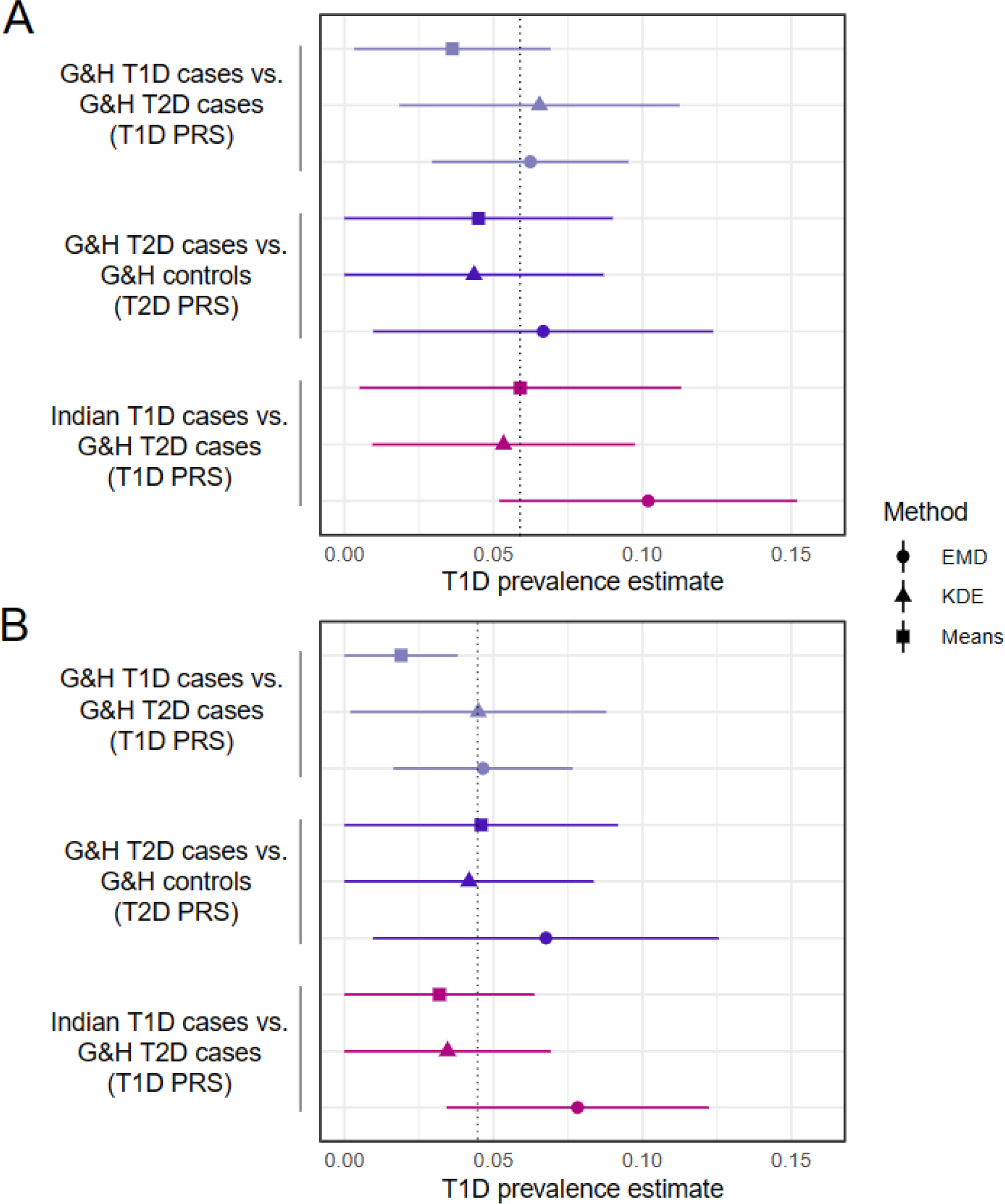
Estimated prevalence of T1D in the G&H ambiguous group. Panel (A) includes all individuals in the ambiguous group (n= 839) and (B) excludes those individuals where only a T1D clinical code is present (n = 821). Points show mean estimates and horizontal lines indicate 95% confidence intervals. The dotted line indicates the median point estimate across all methods. The point type indicates the statistical method used for estimation. EMD: Earth Mover’s Distance; KDE: Kernel density estimation.

### Associations between PRSs and clinical characteristics in G&H

Figure 2 shows the average T1D and T2D PRSs in the four clinically-defined groups in G&H. The ambiguous group had a similar (two-tailed Wilcoxon signed-rank tests, *P*=0.39) T1D PRS to the T2D group, which is consistent with it only containing a small proportion of T1D cases. The ambiguous group has a greater T2D PRS than the T2D group (*P*=5.6×10^-4^), likely because the ambiguous cases were defined as having earlier age of onset, which has previously been shown to be associated with higher polygenic risk score in T2D cases (37,38). We used multiple linear regression to assess the association between the T1D and T2D PRSs and clinical features (age at diagnosis, BMI at diagnosis, time to insulin, and the presence of T1D and T2D diagnostic code) in the ambiguous group and the reference T2D group (Figure 3). Age of diagnosis was significantly negatively associated with the T2D PRS within the T2D cases (*P*=3.73×10^-7^), as expected (37,38). There was no significant association between BMI at diagnosis, time to insulin or the presence of T1D/T2D diagnostic codes with any of the PRSs.

**Figure 2.**
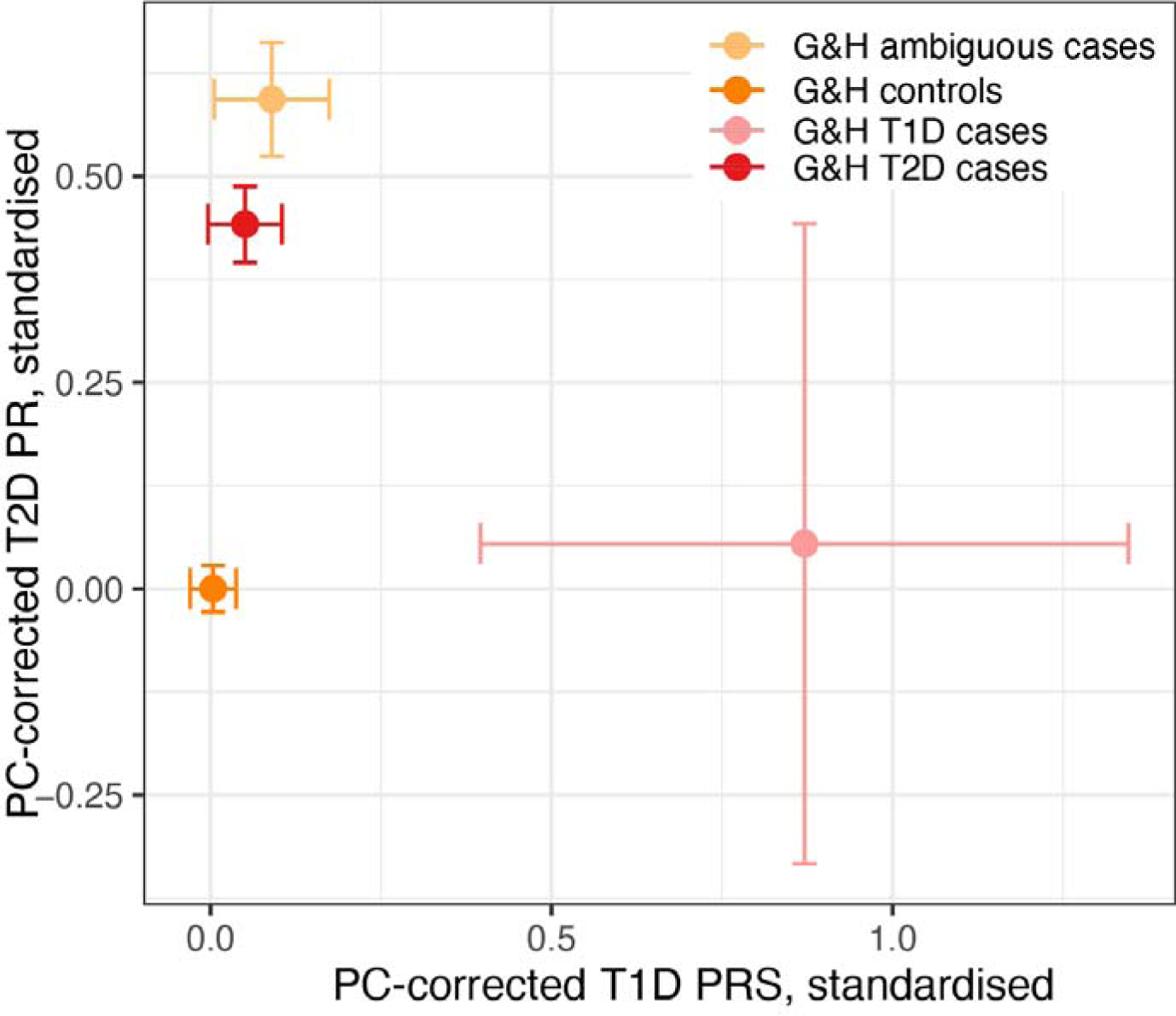
Average PC-corrected T1D and T2D polygenic risk score (PRS) of subgroups in Genes and Health (G&H), with 95% confidence intervals. These have been standardised such that the controls have a mean of 0 and variance 1.

**Figure 3.**
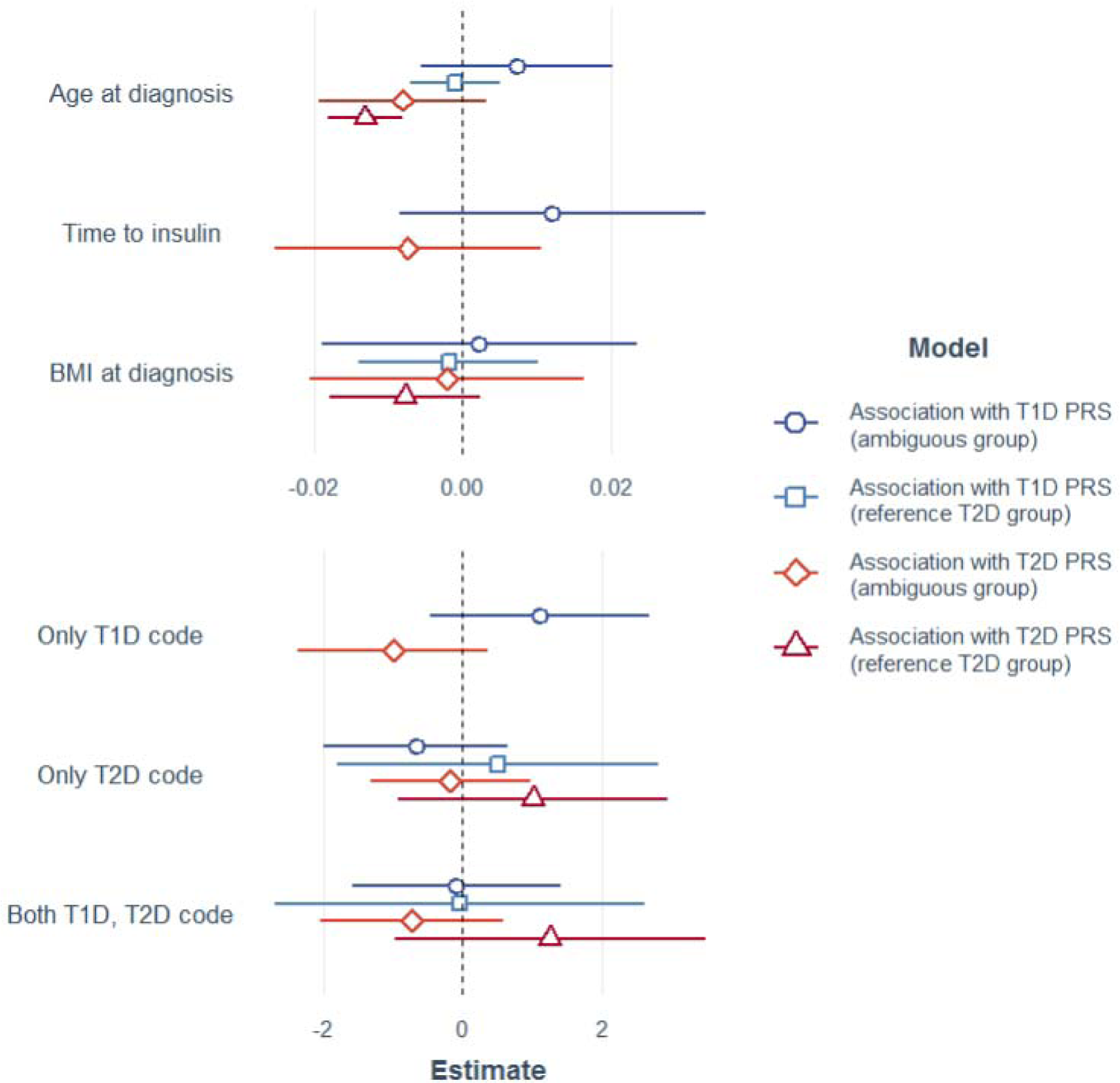
Results from multiple linear regressions of the PC-corrected T1D or T2D PRSs on the indicated clinical variables within either the ambiguous or T2D cases from G&H. Points show the point estimates for the effect size and lines show the 95% confidence intervals. The estimates are split into two panels due to the difference in their scale. Note that the regression within the reference T2D group excluded ‘time to insulin’ and ‘having only a T1D code’ since these were not relevant because of how this group was defined.

## Discussion

Our study combines routine health data from a large population-based study of British Bangladeshi and Pakistani individuals with ancestry-optimised polygenic risk scores to estimate the proportion of T1D cases, and the rate of misclassification, in a group of people with diabetes who have ambiguous clinical features. We undertook this study due to the well-recognised challenges of correctly classifying diabetes type in South Asian populations in which young onset T2D is increasingly common and differences in fat distribution and fat mass mean clinical features such as BMI are unhelpful. Whilst similar methods have previously been employed to aid diabetes diagnosis (25), this study is the first to systematically assess the likely rates of misclassification in a large, real world south Asian population receiving routine diabetes care.

We used standard criteria to define T1D and T2D cases from our population-based cohort with linked health record data. Diabetes autoantibodies and measures of beta cell function (serum C-peptide) were rarely recorded. We derived a large group of individuals whose diabetes diagnosis was clinically ‘ambiguous’ but was characterised by insulin treatment. The clinically ambiguous group included people classified (by diagnostic codes) as having T1D, T2D or both. Using data from an Indian T1D reference cohort and ancestry-optimised PRSs corrected for population structure, we estimated that the true proportion of T1D in this clinically ambiguous group is most likely in the range of 3.6-10.2% (median estimate across approaches = 5.9%, although with wide 95% confidence intervals). Diagnostic codes were not significantly associated with the T1D and T2D PRSs within the ambiguous group (Figure 3), suggesting misclassification is therefore likely. When we removed those individuals who had only diagnostic codes for T1D (i.e. those where we assume clinical suspicion for T1D was highest) from the analysis, and re-estimated the proportion of T1D cases amongst the remaining ambiguous individuals, we obtained a median estimate of 4.5% across approaches, which we regard as an estimate of putative misclassification rate. This is lower than estimates obtained in previous studies in Europeans that only relied on broad clinical criteria (7-15% (6,7), but it is difficult to draw conclusions from this due to our wide error bars, as well as the distinct populations, cohorts, and methodologies. These published estimates based on clinical criteria may be over-estimates of the true misclassification rate.

Our work builds on the methods developed by Evans et al (22), and applies ancestry-adjusted polygenic scores to estimate disease prevalence within clinically-defined groups of individuals with diabetes. This approach allowed us to employ a set of reference cases with different recent genetic ancestry to the target population (i.e. Indian versus Pakistani/Bangladeshi). We conducted joint PCA of both the reference and target sample combined, to ensure that the PCs captured the genetic diversity within the full sample, particularly given the strong fine-scale population structure in South Asia (39,40). It was critical to carry out the ancestry correction using the relationship between the PCs and PRSs defined in controls alone, since, within our sample, T1D case status was strongly correlated with ancestry and hence the PCs, and we did not wish to ‘correct away’ the true difference in T1D PRS between cases and controls.

Our study has certain limitations. The low numbers of T1D cases that could be robustly identified by clinical criteria in our G&H sample meant that we could not optimise the weights and choice of SNPs for the T1D PRS within this sample, or use the true T1D cases from G&H as our reference sample when estimating the fraction of T1D cases in the ambiguous group. The steps outlined above using T1D cases from a study of Indian individuals and a PCA-based method to correct for population structure has mitigated this. If we had access to a well-powered T1D GWAS within individuals of Pakistani and Bangladeshi ancestry, this would likely boost the accuracy of the PRSs and improve the accuracy of our inference. The use of routine health data in our analyses has potential limitations: specifically, we found very low numbers of individuals who had recorded diabetes autoantibodies and measures of beta cell function (C-peptide) that are used to classify diabetes type. This observation could reflect missing (likely historic) data and lead to the inclusion of true T1D cases in our ambiguous group and overestimation of misclassification. However, the absence of these test results in the multiple data linkages we used is likely to mirror the reality of data availability in clinical care (either due to tests not being performed, or being performed in a different healthcare setting from which data are not available) and the diagnostic uncertainty arising from this. We showed that the clinical features that are commonly used to help classify T1D and T2D (age and BMI at diagnosis, and time to insulin) were not associated with the ancestry-optimised PRS within the ambiguous group, implying that they have limited utility to help distinguish T1D from T2D within this group. However, we were unable to analyse data on diabetic ketoacidosis, a clinical feature that can also aid diagnostic classification, due to the low frequency (and presumed poor quality) of diagnostic codes in our data linkage. Whilst we have excluded individuals with diagnostic codes for rare types of diabetes (monogenic, secondary), it is possible that there are individuals with undiagnosed monogenic diabetes in our ambiguous group who have not been excluded and who could affect our estimate of the proportion of T1D. Finally, our study does not attempt to estimate the total burden of misclassification. Rather, we restricted our analysis to a group of individuals with diabetes and ambiguous clinical features who were treated with insulin, and excluded those who were not insulin treated and were within three years of their diabetes diagnosis (i.e. those who did not meet the T2D clinical criteria). By restricting our analyses to those who are insulin-treated, we have identified an important subpopulation of people with diabetes who can be readily identified through clinical systems and targeted for further diagnostic assessment (e.g. with measurement of C-peptide and diabetes autoantibodies) to assist correct classification.

Our study confirms that correct classification of diabetes is difficult in populations of British South Asians, and that routinely recorded clinical features at diagnosis do not discriminate between type 1 and type 2 diabetes. Our PRS-determined estimates of T1D prevalence and misclassification demonstrate that within the ambiguous group, one in twenty individuals have not been correctly identified as having T1D. This is an important finding as it suggests that many individuals are receiving poorly-targeted clinical care, are at greater risk of hospital admission due to diabetic emergencies, and may be missing out on technology-supported care such as insulin pump therapy. Such ‘ambiguous’ diabetes cases could be readily identified in primary care settings by routinely collected health data. We propose that T1D could be identified robustly in the majority of these ambiguous cases using diabetes autoantibody and C-peptide measurements, which are currently performed infrequently. Should polygenic risk scores be implemented in routine clinical care in the future (41), these could be readily included in diagnostic tests to aid the correct classification of type 1 and type 2 diabetes, but only with careful optimisation for different ancestry groups.

## Data Availability

The Genes & Health data are available upon application to the Genes & Health Executive Committee, as outlined here https://www.genesandhealth.org/research/scientists-using-genes-health-scientific-research. Access will be granted within a trusted research environment, as required by the ethical approvals. Request for access to the Indian cohort should be sent to Giriraj R Chandak, MD at.

## Acknowledgments

**For the Genes & Health cohort**

We thank Social Action for Health, Centre of The Cell, members of our Community Advisory Group, and staff who have recruited and collected data from volunteers. We thank the NIHR National Biosample Centre (UK Biocentre), the Social Genetic & Developmental Psychiatry Centre (King’s College London), Wellcome Sanger Institute, and Broad Institute for sample processing, genotyping, sequencing and variant annotation. We thank: Barts Health NHS Trust, NHS Clinical Commissioning Groups (City and Hackney, Waltham Forest, Tower Hamlets, Newham, Redbridge, Havering, Barking and Dagenham), East London NHS Foundation Trust, Bradford Teaching Hospitals NHS Foundation Trust, Public Health England (especially David Wyllie), Discovery Data Service/Endeavour Health Charitable Trust (especially David Stables), and NHS Digital for GDPR-compliant data sharing backed by individual written informed consent.

Most of all we thank all of the volunteers participating in Genes & Health and UK Biobank.

We thank the current Genes & Health Research Team (in alphabetical order by surname): Shaheen Akhtar, Mohammad Anwar, Omar Asgar, Samina Ashraf, Saeed Bidi, Gerome Breen, James Broster, Raymond Chung, David Collier, Charles J Curtis, Shabana Chaudhary, Megan Clinch, Grainne Colligan, Panos Deloukas, Ceri Durham, Faiza Durrani, Fabiola Eto, Sarah Finer, Joseph Gafton, Ana Angel Garcia, Chris Griffiths, Joanne Harvey, Teng Heng, Sam Hodgson, Qin Qin Huang, Matt Hurles, Karen A Hunt, Shapna Hussain, Kamrul Islam, Vivek Iyer, Ben Jacobs, Ahsan Khan, Cath Lavery, Sang Hyuck Lee, Robin Lerner, Daniel MacArthur, Daniel Malawsky, Hilary Martin, Dan Mason, Rohini Mathur, Mohammed Bodrul Mazid, John McDermott, Caroline Morton, Bill Newman, Elizabeth Owor, Asma Qureshi, Samiha Rahman, Shwetha Ramachandrappa, Mehru Reza, Jessry Russell, Nishat Safa, Miriam Samuel, Michael Simpson, John Solly, Marie Spreckley. Daniel Stow, Michael Taylor, Richard C Trembath, Karen Tricker, Nasir Uddin, David A van Heel, Klaudia Walter, Caroline Winckley, Suzanne Wood, John Wright, Julia Zollner.

## Authors’ relationships and activities

SF and HCM have received salary contributions via the Genes & Health Industry Consortium of AstraZeneca PLC, Bristol-Myers Squibb Company, GlaxoSmithKline Research and Development Limited, Maze Therapeutics Inc, Merck Sharp & Dohme LLC, Novo Nordisk A/S, Pfizer Inc, Takeda Development Centre Americas Inc. ML has received speaker’s fees from Medtronic and Insulet.

## Author Contributions

SF, HCM, and GP conceived the project. GP prepared, cleaned and analysed the clinical data and defined the clinical cohorts, with contribution from SH and ML. TL and THH undertook QC of the Genes & Health genotype data. AS generated the genotype data from the Indian cohort, and carried out initial QC on these, in consultation with GRC. TL undertook QC of Genes & Health and further QC on the Indian genotype data and imputed both datasets to TopMED with input from AS, DF, MW, RO and GRC. TL conducted the PCA and calculated the PRSs with input from AS, QQH, RO, MW, and GRC. TL correlated the PRSs with clinical data and, with help from NT, ran the estimation method. DvH and SF lead the G&H study and, with the G&H Research team, they supervise all data collection. CSY created the Indian clinical cohort and GRC supervised genotyping of its cases and controls with AS. Intellectual contribution to analyses was provided by RO, MW, NT, CSY and GRC. SF, HM, GP and TL wrote the first draft of the manuscript, and all authors commented on it.

## Data availability

The Genes & Health data are available upon application as described here https://www.genesandhealth.org/research/scientists-using-genes-health-scientific-research. Access will be granted within a custom-built trusted research environment. Request for access to the Indian cohort should be sent to Giriraj R Chandak at and Chittaranjan Yajnik.

## Electronic supplementary material

### ESM Methods

#### Definition of clinical diagnoses in Genes & Health

We captured all diagnostic codes for type 1 (T1D) and type 2 diabetes (T2D) from primary care (SNOMED) and secondary care (ICD-10), using clinical codesets in routine use in the NHS. These have been previously used (21) and are summarised in ESM Table 1. We excluded all individuals with diagnosed secondary diabetes, rare types of diabetes (e.g. monogenic diabetes) or a history of pancreatitis or pancreatic surgery from all analyses using additional codelists (see ESM Table 1). We collated additional clinical features, including prescribing data, clinical measurements and laboratory data, including measurements of body mass index (BMI), Haemoglobin A1c (HbA1c), high-density lipoprotein (HDL) and triglycerides (TG) measured within a year of the earliest date of diagnosis. Where C-peptide measures were available (either fasting or random samples), we defined insulin deficiency as C-peptide <200 pmol/l (measured fasting, random or stimulated). We defined diabetes autoantibody positivity as the presence of any NHS laboratory-defined anti-glutamic acid, anti-ZnT8 or anti-islet cell antibody titre above the local laboratory reference range. We calculated time to first use of insulin for patients who had received insulin based on prescription records and dates of diagnosis. We also captured the diagnosis of other organ-specific autoimmune diseases (ESM Table 2).

#### Cleaning of quantitative data from G&H electronic health records

BMI at diagnosis was calculated based on the weight nearest to diagnosis date and the median adult height for patients diagnosed over age 18, with clinically improbable results excluded (BMI >75 or <12 kg/m^2^). Laboratory results were considered outliers if they fell out with the established reference range for our local NHS laboratory (HbA1c > 160 mmol/l, HDL > 6.24 mmol/l, triglycerides > 50.0 mmol/l). For older HbA1c results, values expressed in Diabetes Control and Complications Trial percentage units were converted to mmol/mol using an established conversion formula.

#### Preparation of the genotype data from Genes & Health

##### Quality control

We performed quality control of the Global Screening Array data with Illumina’s GenomeStudio and plink v1.9. Using GenomeStudio, we removed variants with h-cluster separation scores <0.57, Gentrain score <0.7, an excess of heterozygotes >0.03, or ChiTest 100 (Hardy-Weinberg test) <0.6. We additionally removed variants that were included on the array in order to tag specific structural variants, and samples with that failed gender checks or had low call rate (<0.995 for male samples and <0.992 for female samples across all 637,829 variants including those on Y chromosome for males). We then removed SNPs with MAF < 0.5%, leaving 44,396 people and 399,911 SNPs. To ensure we did not lose SNPs that have high quality but that were out of Hardy-Weinberg Equilibrium due to high rates of consanguinity and strong population structure in Pakistanis (40), we removed SNPs that failed Hardy-Weinberg Equilibrium p-value < 1×10^-6^ in Bangladeshis alone, as done in (28). This left 399,047 SNPs.

##### Inference of genetic ancestry

To infer genetic ancestry, we merged the SNPs with MAF>1% from the imputation dataset (355,136 SNPs) with whole-genome sequence data from unrelated individuals (inferred using KING as described below) from the 1000 Genomes Project and with Central and South Asian individuals from the Human Genome Diversity Project. After first excluding palindromic variants and multiallelic sites from both datasets, we merged the datasets by matching positions and alleles of the common SNPs that passed QC in G&H, and kept variants found in both datasets, leaving 349,632 SNPs. We then excluded 1,285 variants due to allele frequency discrepancies between G&H and South Asian reference individuals (>4 standard deviations from the mean residual of -log_10_ frequency bins, and Fisher’s exact test P<1×10^-5^), leaving in 348,347 SNPs. Before carrying out the principal components analysis (PCA), we pruned the SNPs (window size 1000kb, step size 50, linkage disequilibrium (LD) r^2^ cutoff 0.1) and removed long LD regions, leaving 104,552 SNPs. We ran the PCA with plink 1.9, first calculating PCs for the 3,433 reference individuals, then projecting the G&H samples into the reference PC space. We used the umap package in R to perform UMAP on the PCs, and found that the reference individuals separated into known superpopulations optimally when using seven PCs. This allowed us to remove 76 of the G&H individuals who were genetically most similar to non-South Asian samples from the reference datasets leaving 44,320 individuals. We then performed a second PCA on the unrelated G&H individuals, projecting the related G&H individuals on those, and then performing UMAP with four PCs to identify two major clusters. These corresponded well to self-identified Pakistani and Bangladeshi groups, and were thus used to define individuals as genetically Pakistani or Bangladeshi.

##### Inference of relatedness

We used the PropIBD metric from KING to estimate relatedness between the 9,675 genetically-inferred Pakistani or Bangladeshi individuals in the following groups, listed here in priority order: 1. T1D cases, 2. Ambiguous cases, 3. T2D cases and 4. non-diabetic controls. We then removed one from each pair of related individuals inferred to be third-degree relatives or closer, preferentially retaining individuals in the higher priority groups. To maximise the sample size, we first removed the relatives within the group of interest (starting with T1D cases, followed by the other groups in priority order). We then ranked individuals by their number of inferred relatives, then sequentially removed the individual with the highest number of relatives from the groups of lower priority, until no relative pairs remained.

#### Quality control of genotype chip data in the Indian cohort

Genotype data on the Indian subjects were generated using the Global Screening Array v3.0. Genome studio 2.0 was used to convert microarray intensity files (.idat) to genotypes. We applied a sample call rate of 95% and Gentrain > 0.5, cluster separation > 0.4 and AA/AB/BB Tdev < 0.06 for the SNPs. Further poorly clustering SNPs were manually inspected and removed as per recommendations (42). This retained 649 people and 594,542 SNPs. Using plink v1.9, we applied MAF < 0.5% and Hardy-Weinberg p-value < 1×10^-6^, leaving 341,343 SNPs. The data were merged with QCed data from the G&H cohort before imputation, as described in the main Methods.

#### Principal component analysis and imputation of G&H and Indian samples

We first merged the post-QC genotype data from the G&H and Indian cohorts, then pruned the SNPs (window size 1000kb, step size 50, LD r^2^ cutoff 0.1) and removed regions of long-range linkage disequilibrium except those overlapping the HLA region (chr6:25391792-33424245), leaving 113,928 SNPs. We then carried out PCA using plink v1.9.

Imputation was carried out using the Michigan TOPMED Imputation Server on the common subset of genotyped variants between the G&H and Indian samples, excluding palindromic SNPs and chrY SNPs. Non-ACGT variants were removed using --snp-only just-acgt in plink2 at a MAF cut-off of 0.005. TOPMed r2 was used as the imputation reference panel. We removed variants with imputation accuracy r^2^<0.3, leaving 45,946,210 SNPs.

### ESM Tables

**ESM Table 1:** Clinical codelists used to define the set of type 1 and type 2 diabetes cases. Codes included in the NHS primary care Quality and Outcomes Framework are denoted in blue.

**ESM Table 2:** Clinical code lists used to define autoimmune diseases.

**ESM Table 3:** Clinical codes denoting people at risk of diabetes, which were used to exclude people from the control group. Codes included in the NHS Primary Care Quality and Outcomes Framework are denoted in blue.

**ESM Table 4.** Numbers of samples remaining after each filter used to define T1D, T2D and ambiguous cases and the controls in G&H.

**ESM Table 5:** Exact p-values for post-hoc comparisons of clinical features between the groups in G&H. The group with which the comparison was made is indicated before the p-value. If the post-hoc comparison was not made (e.g. there was no statistical significance at ANOVA or if only two groups were compared for this feature), NA is given.

### ESM Figures

**ESM Figure 1.**
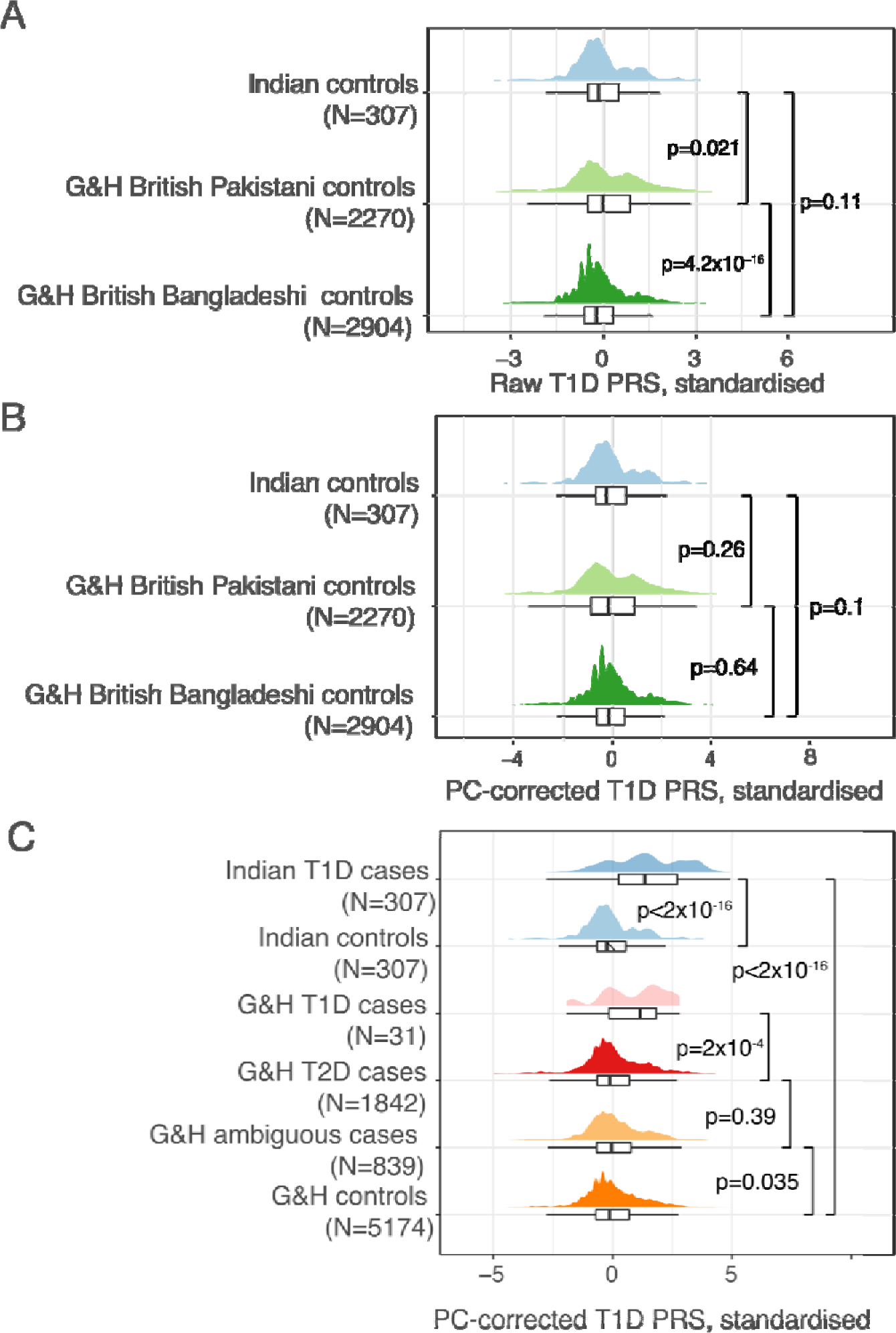
Distribution of T1D polygenic risk scores (PRS) in the Indian and Genes and Health (G&H) clinical groups. A) and B) show the distributions in the control groups split by ancestry, before and after PC correction respectively. C) shows the distributions in the various groups of cases and controls used in the analyses, after PC-correction. P-values are from Wilcoxon signed-rank tests comparing the two indicated groups. ESM Figure 4 shows the equivalent distributions for the T2D PRS.

**ESM Figure 2.**
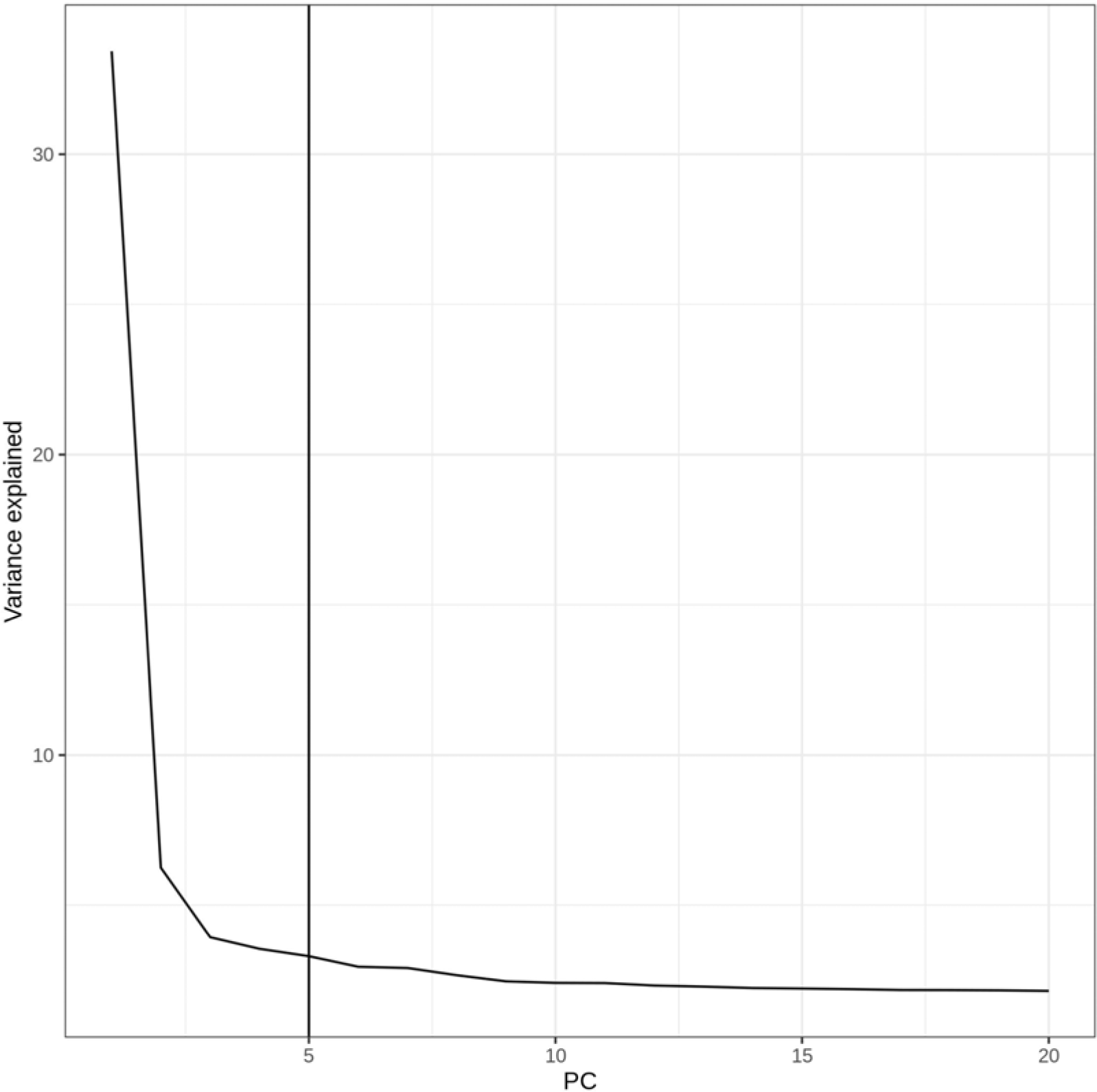
Scree plot indicating the variance explained by principal components 1-20 from the PCA of G&H and Indian samples. Based on this, we decided to regress out five principal components from the PRSs.

**ESM Figure 3.**
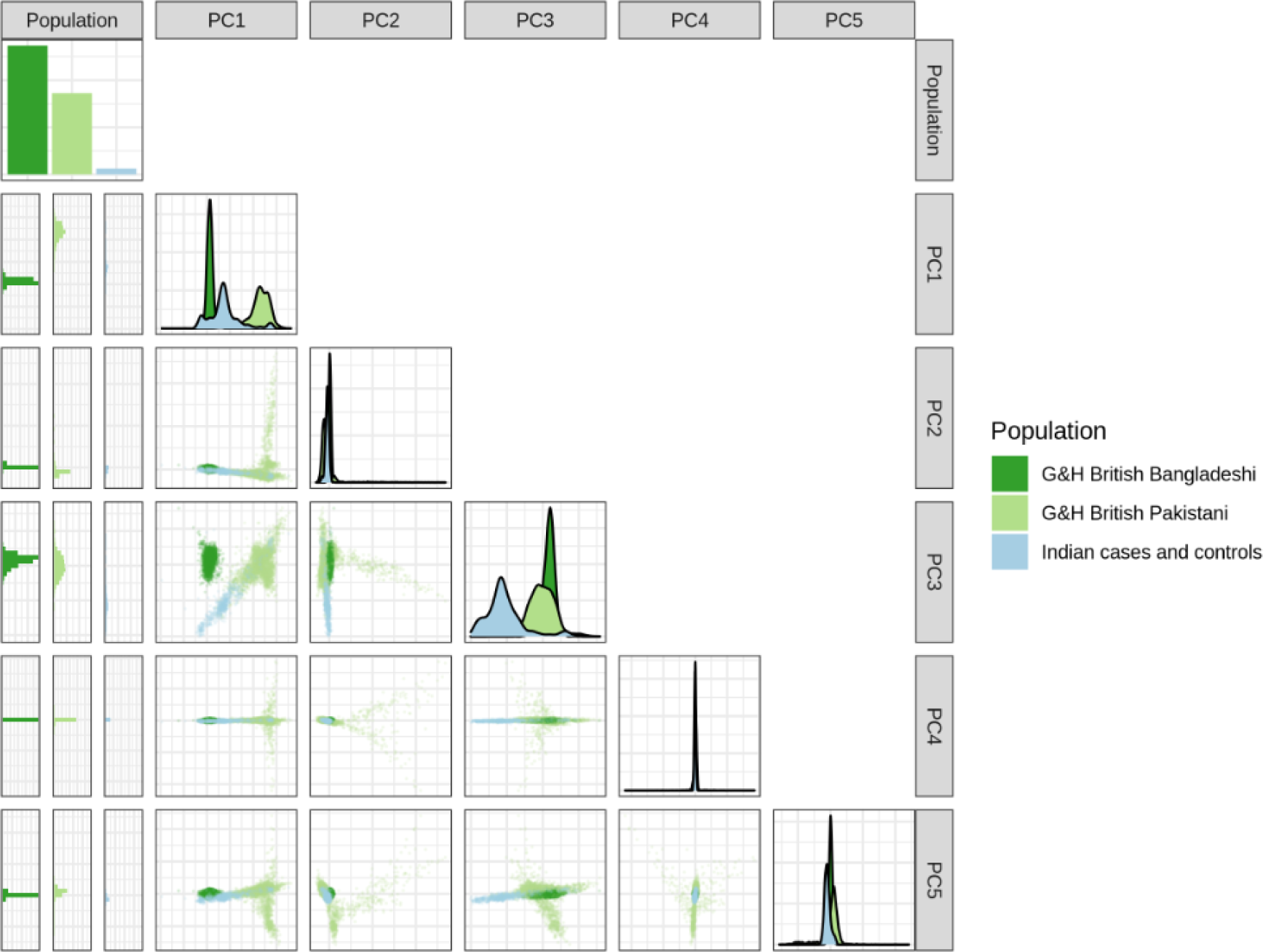
Principal component analysis of G&H and Indian samples (cases and controls combined).

**ESM Figure 4.**
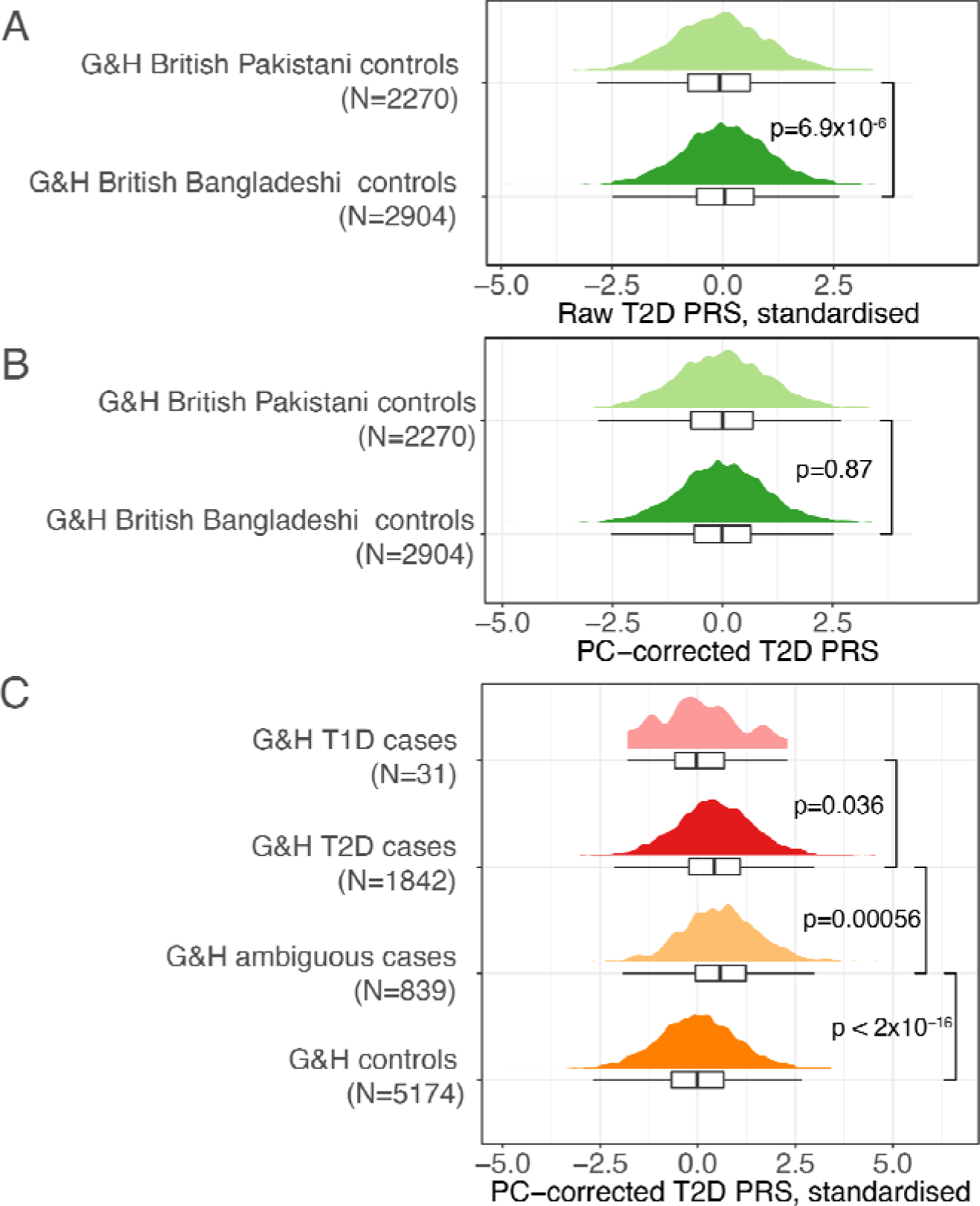
Distribution of T2D polygenic risk scores (PRS) in the G&H clinical groups. A) and B) show the distributions in the control groups split by ancestry, before and after PC correction respectively. C) shows the distributions in the various groups of cases and controls used in the analyses. P-values are from Wilcoxon signed-rank tests comparing the two indicated groups.

